# Temperature at conception and pregnancy loss in rural KwaZulu-Natal Province, South Africa: Implications for climate change policy in sub-Saharan African settings

**DOI:** 10.1101/2021.03.18.21253882

**Authors:** Yoshan Moodley, Frank Tanser, Andrew Tomita

## Abstract

**Background:** Global warming is projected to cause a substantial rise in temperatures with serious health implications across sub-Saharan Africa. Although South African policy makers have drafted a climate change adaptation plan, potential health threats posed by increasing temperatures on women’s reproductive health are overlooked due to the lack of local population-based evidence. We sought to address the gap in the evidence around global warming and women’s reproductive health in sub-Saharan Africa by using one of the continent’s largest prospective cohorts from rural KwaZulu-Natal Province, South Africa to investigate the relationship between temperature at conception and pregnancy loss.

**Methods:** Our study sample consisted of 36341 pregnancies from 16765 women from the uMkhanyakude District of KwaZulu-Natal, South Africa between 2000-2017. Average monthly temperatures for the study locale during the study period were obtained from the South African Weather Services. An adjusted logistic regression model was used to investigate the relationship between temperature at conception and pregnancy loss (miscarriage or stillbirth).

**Results:** The rate of pregnancy loss in the study sample was 1.9 (95% Confidence interval [CI]: 1.7-2.0) per 100 pregnancies. We observed a 4% higher odds of pregnancy loss for each 1°C increase in temperature (Adjusted Odds Ratio: 1.04, 95% CI: 1.01-1.08).

**Conclusion:** There is a clear relationship between temperature and pregnancy loss in our sub-Saharan African setting. The effects of global warming will likely exacerbate the existing challenges for women’s reproductive health in this region. Pregnancy outcomes should be given adequate attention when sub-Saharan African governments draft policies in response to global warming.

## 1. Introduction

Despite the international commitments made as part of the landmark United Nations Framework Convention on Climate Change (UNFCCC) Paris Agreement to mitigate the impact of global warming on human health, most sub-Saharan African countries who have ratified the Agreement will struggle to restrict rises in national ambient temperatures below the proposed target of 1.5°C (Weber et al., 2018). Sub-Saharan Africa is particularly vulnerable to the hazardous effects of global warming which will likely exacerbate existing high ambient temperatures in the region. In addition, resource constraints are anticipated to hinder the effective roll-out of adaptation or mitigation strategies in sub-Saharan Africa (Weber et al., 2018; Russo et al., 2016).

Global warming is projected to cause a 6°C rise in maximum temperatures across South Africa by the year 2070 (Mbokodo et al., 2020), which is expected to have dire implications for the health of socially vulnerable populations in parts of the country that will likely still be reeling from the aftermath of a stubborn HIV epidemic. Several South African studies have reported an association between high ambient temperatures and an increased risk of adverse health outcomes such as mortality, heat stress, and infectious diseases (Chersich et al., 2018). South African policy makers have acknowledged this setting-specific evidence and proactively drafted a climate change adaptation plan in response to global warming and the impending increase in national maximum temperatures (Garland, 2014). Regrettably, women’s reproductive health is overlooked in this plan due to the lack of local population-based evidence (Garland, 2014). With excessively high poverty, HIV infection, and maternal mortality rates (Ngwenya et al., 2018; Vandormael et al., 2019; Tlou et al., 2017), sub-Saharan African women of childbearing age are a prime example of an already vulnerable population which could potentially suffer additional adverse health outcomes due to global warming. Although the relationship between ambient temperature and fetal health is still to be established for sub-Saharan African settings, reports from more industrialized regions of the world suggest a relationship between higher ambient temperatures and adverse pregnancy outcomes such as preterm delivery, low birth weight, and pregnancy loss (Kuehn and McCormick, 2017; Bekkar et al., 2020).

We sought to address the gap in the evidence around global warming and women’s reproductive health in sub-Saharan Africa by using one of the African continent’s largest prospective cohorts from rural KwaZulu-Natal, South Africa to investigate the relationship between temperature at conception and pregnancy loss.

## 2. Methods

### 2.1 Study design and setting

This was a population-based, prospective cohort study involving pregnant women from a rural community in the uMkhanyakude District of KwaZulu-Natal, South Africa. The rural community, described in detail elsewhere (Tanser et al., 2008), has been part of the Africa Health Research Institute’s (AHRI’s) health and demographic surveillance systems for over 20 years. The uMkhanyakude District has a high unemployment rate and relatively poor levels of service delivery (Mulopo et al., 2020). The two most important public health problems facing women of childbearing age in the uMkhanyakude District include a high HIV incidence of 3.06 seroconversion events per 100 person-years and a high maternal mortality ratio of 650 maternal deaths per 100000 live births (Vandormael et al., 2019; Tlou et al., 2017). As shown in Fig. 1, the uMkhanyakude District is located in a geographic region with a sub-tropical climate characterized by wet summers and dry winters (Morgenthal et al., 2006). Daily maximum temperatures in the uMkhanyakude District are usually above 20°C (Morgenthal et al., 2006).

**Fig. 1.**
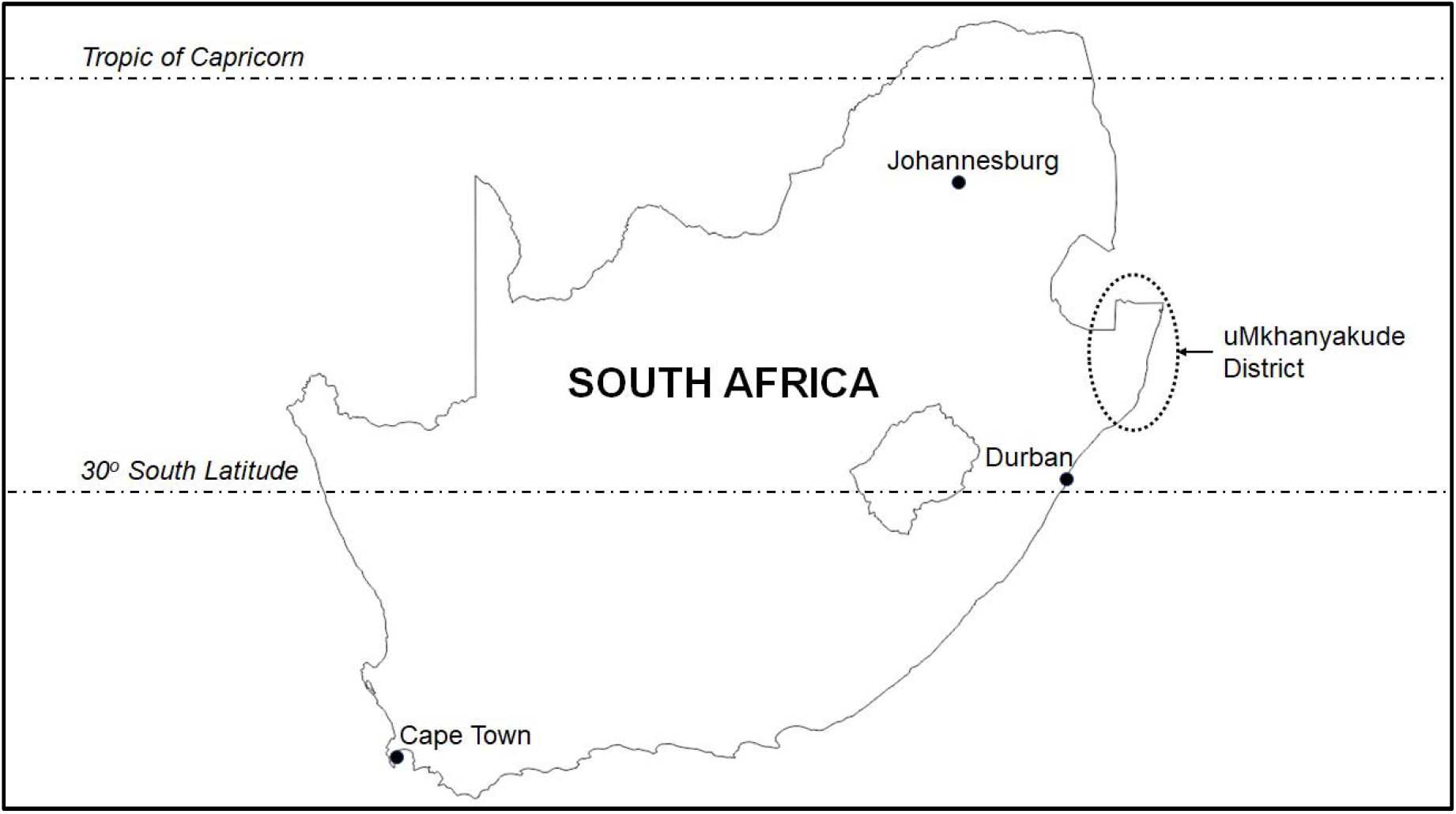
Location of the uMkhanyakude District, South Africa

### 2.2 Data sources

AHRI has conducted regular population-based surveys in the rural community since 2000. These surveys incorporate a pregnancy questionnaire that is administered (biannual until 2011, triannual since 2012) to all women in the community who report pregnancies (Chetty et al., 2016; Chetty et al., 2017). The pregnancy questionnaire collects data on several maternal and pregnancy-related characteristics such as number of prior pregnancies, date of last menses, antenatal clinic attendance during the pregnancy, pregnancy outcome date, as well as pregnancy outcomes such as live births, number of children delivered, abortions, and stillbirths (Chetty et al., 2017).

Data for other variables which might influence pregnancy outcomes is collected in other demographic and health questionnaires that are also administered during the population-based survey (Tanser et al., 2008). This includes sociodemographic characteristics such as age, socioeconomic status based on a household asset index, marital status, and educational level; and infectious diseases which are endemic to this region of South Africa (HIV and tuberculosis). Temperature data used in the analysis was the average maximum temperature in the study locale for each month between January 2000 and December 2017 (i.e. 216 average monthly measurements over the entire study period), and was obtained from the South African Weather Services.

### 2.3 Measures

The study outcome, pregnancy loss, was defined as a either miscarriage or stillbirth. Miscarriage was defined as a pregnancy of <28 weeks gestational duration (Chetty et al., 2017; Moodley et al., 2021). This was established for each pregnancy by calculating the difference in weeks between the date that the pregnancy ended and the date of the last menses. Stillbirth was collected as one of the pregnancy outcomes in the pregnancy questionnaire and was defined as delivery of a dead infant of gestational age ≥28 weeks (Chetty et al., 2017; Moodley et al., 2021). The date of conception was determined using the date of the last menses. Temperature at conception was defined as the temperature corresponding to the month and year in which the pregnancy was conceived.

### 2.4 Eligibility criteria

We reviewed records from AHRI population-based surveys conducted between 2000 and 2017 for all singleton pregnancies conceived in women aged 16-35 years old. We excluded pregnancies that were electively aborted. Our study sample was comprised of 36341 pregnancies from 16765 women.

### 2.5 Statistical analysis

R version 3.6.2 (R Foundation for Statistical Computing, Vienna, Austria) was used to analyze the study data. We used descriptive statistics to summarize the distribution of various pregnancy-related and maternal characteristics in our study sample. We then performed univariate and multivariate logistic regression analyses to investigate the relationship between temperature at conception and pregnancy loss in our study sample. The results of our univariate and multivariate regression analyses are presented as unadjusted and adjusted odds ratios (ORs), with 95% confidence intervals (95% CI).

### 2.6 Ethical approval

AHRI’s active health and demographic surveillance platform, which includes the population-based cohort from which our study data was obtained, has been approved by the Biomedical Research Ethics Committee of the University of KwaZulu-Natal, South Africa (Protocol reference: BE290/16). All participants in the AHRI population-based surveys provided written informed consent.

### 2.7 Availability of data and materials

The datasets analyzed in this study are available on AHRI’s Online Data Repository [https://data.ahri.org/index.php/home].

## 3. Results

The study sample (N=36341) is described in Table 1. The rate of pregnancy loss in the study sample was 1.9 (95% CI: 1.7-2.0) per 100 pregnancies. Of the 679 pregnancies that were lost, 395 (58.2%) were miscarriages and the remaining 284 (41.8%) were stillbirths. The average maternal age for pregnancies comprising the sample was 23.74 (95% CI: 23.69-23.79) years. A prior history of pregnancy loss was rare (n=979, 2.7%). A maternal history of tuberculosis was also rare (n=226, 0.6%). Two-thirds of all pregnancies were in mothers from the low socioeconomic status group (n=24444, 67.3%). Only a small percentage of women complied with the national recommendation of attending at least eight routine antenatal clinic visits during their pregnancy (n=522, 1.4%). A large proportion of pregnancies were in unmarried mothers (n=34662, 94.8%). Half of all pregnancies were in mothers who had already completed their primary school education (n=19117, 52.6%). Lastly, 2911 pregnancies (8.0%) occurred in mothers who were HIV-positive at the time.

**Table 1.**
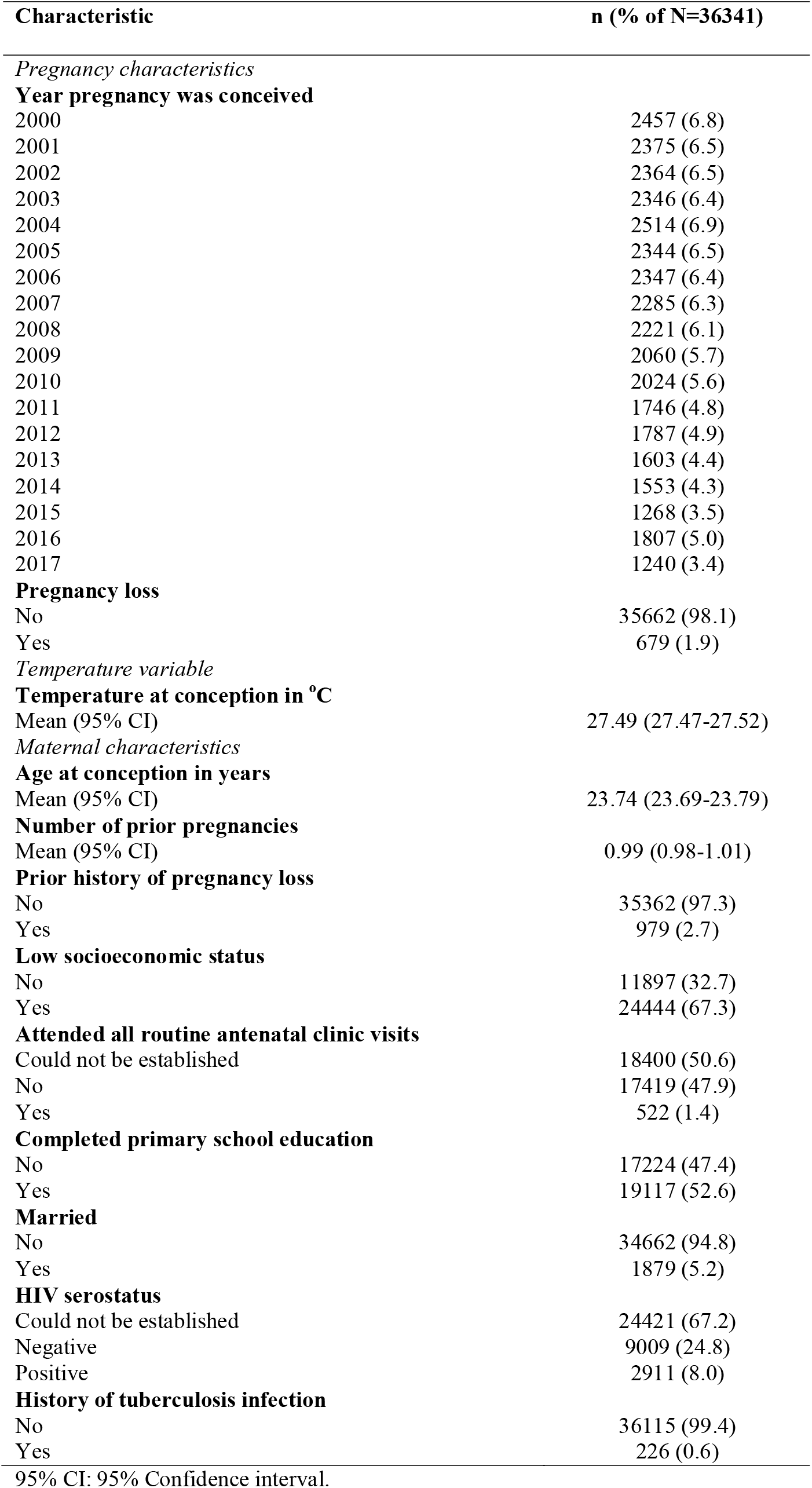
Description of the study sample

Table 2 presents the results of the univariate and multivariate logistic regression analyses. Our univariate statistical analysis revealed a 5% higher odds of pregnancy loss for each 1°C increase in temperature at conception (unadjusted OR: 1.05, 95% CI: 1.02-1.09). When we adjusted our analysis for potential confounders, we observed a 4% higher odds of pregnancy loss for each 1°C increase in temperature at conception (adjusted odds ratio: 1.04, 95% CI: 1.01-1.08).

**Table 2.** Results of the univariate and multivariate logistic regression analyses investigating the relationship between temperature at conception, various covariates, and pregnancy loss

| Characteristic | Unadjusted OR (95% CI) | Adjusted OR (95% CI) |
| --- | --- | --- |
| *Temperature variable* |  |  |
| **Temperature at conception (per 1°C increase)** | 1.05 (1.02-1.09) | 1.04 (1.01-1.08) |
| *Maternal characteristics* |  |  |
| **Age at conception (per 1-year increase)** | 1.01 (1.00-1.03) | 1.01 (0.99-1.03) |
| **Number of prior pregnancies (per additional pregnancy)** | 1.05 (0.99-1.11) | 0.47 (0.41-0.53) |
| **Prior history of pregnancy loss** |  |  |
| No | 1.00 | 1.00 |
| Yes | 69.98 (58.97-83.04) | 226.93 (176.64-291.53) |
| **Low socioeconomic status** |  |  |
| No | 1.00 | 1.00 |
| Yes | 1.21 (1.03-1.43) | 1.05 (0.85-1.31) |
| **Attended all routine antenatal clinic visits** |  |  |
| Could not be established | 1.00 | 1.00 |
| No | 1.64 (1.40-1.91) | 1.74 (1.44-2.10) |
| Yes | 0.66 (0.27-1.60) | 0.59 (0.22-1.60) |
| **Completed primary school education** |  |  |
| No | 1.00 | 1.00 |
| Yes | 0.88 (0.76-1.03) | 0.99 (0.81-1.20) |
| **Married** |  |  |
| No | 1.00 | 1.00 |
| Yes | 0.94 (0.66-1.33) | 1.04 (0.67-1.61) |
| **HIV serostatus** |  |  |
| Could not be established | 1.00 | 1.00 |
| Negative | 0.79 (0.65-0.95) | 0.93 (0.73-1.18) |
| Positive | 0.88 (0.66-1.18) | 0.91 (0.63-1.31) |
| **History of tuberculosis infection** |  |  |
| No | 1.00 | 1.00 |
| Yes | 1.69 (0.79-3.59) | 1.74 (0.63-4.80) |

OR: Odds ratio, 95% CI: 95% Confidence interval. Reference category ORs=1.00.

## 4. Discussion

Our analysis of data from one of the largest population-based cohorts on the African continent demonstrates clear relationship between increasing temperature at conception and pregnancy loss. Specifically, our adjusted statistical analysis found that the odds of pregnancy loss increased by 4% for each 1°C increase in temperature at conception. This is, to the best of our knowledge, the first study from sub-Saharan Africa to report a relationship between increasing temperatures and adverse pregnancy outcomes. Furthermore, our findings are in general agreement with the findings of two recent systematic reviews on this subject (Kuehn and McCormick, 2017; Bekkar et al., 2020). Thus, our study findings confirm the importance of climate change and increasing ambient temperatures on adverse pregnancy outcomes in sub-Saharan Africa.

There are currently no South African policies responsive to the potential effects of global warming on women’s reproductive health and pregnancy outcomes, and recommendation of an appropriate policy framework lies beyond the scope of our study. However, our study findings do provide sufficient impetus for an appropriate policy framework which seeks to address the impact of increasing temperatures on women’s reproductive health and pregnancy outcomes to be developed by policymakers in South Africa and other sub-Saharan African countries. In the absence of this policy framework, governments in sub-Saharan Africa should consider enforcing one of the most important human rights enshrined in their national constitutional frameworks and The African Charter on Human and People’s Rights – access to a clean basic water supply (South African Human Rights Commission, 2018). All pregnant mothers in sub-Saharan Africa should have access to clean drinking water to prevent dehydration when ambient temperatures are high (Garland, 2014). Increased maternal water consumption during pregnancy is associated with a significantly lower risk of neural tube defects, oral clefts, musculoskeletal defects, and congenital heart defects (Alman et al., 2017). Although the biological mechanism behind this association is still unknown, it nevertheless suggests that facilitating adequate maternal water consumption early during pregnancy is key to reducing subsequent adverse pregnancy outcomes (Alman et al., 2017). Despite some gains in increasing coverage of piped water access in the uMkhanyakude District, there are some communities where surface water sources continue to be used as a primary water source. For instance, 77.2% of households surveyed in the Madeya Village still use surface water as a primary water source (Mulopo et al., 2020). Barriers to ensuring access to a clean basic water supply in these communities should be investigated and addressed as a matter of urgency. It is tempting to propose the use of electric fans and air conditioning as a heat adaptation strategy for households with pregnant women in sub-Saharan African settings. However, this would be unfeasible for several reasons. The increased electricity demand would require the rollout or upgrading of electric power infrastructure (Farbotko and Waitt, 2011), which would be a very costly undertaking in resource constrained sub-Saharan Africa. In addition, many households in the region might be unable to afford electrification (Ismail and Khembo, 2015). Lastly, air conditioning increases environmentally harmful gas emissions, and this can further contribute toward climate change in the region (Farbotko and Waitt, 2011). As such, there remains a need for future research which seeks to establish appropriate interventions for reducing the impact of high ambient temperatures on women’s reproductive health and pregnancy outcomes in resource limited sub-Saharan Africa.

Our study had several strengths, the most notable of which was our use of data from one of Africa’s largest rural population-based cohorts. We also used consistently measured, accurate temperature data from a regional weather station in our analysis. Lastly, the regular frequency of the AHRI surveys reduced the risk of recall bias around pregnancy outcomes. One of the main limitations of our study was that we were unable to adjust our analysis for the presence of sexually transmitted infections, such as syphilis. Syphilis is associated with an almost 5-fold higher risk of pregnancy loss (Gomez et al., 2013). There may also be a possibility that miscarriage and stillbirth was underreported in the rural community. Furthermore, the observed relationship between temperature and pregnancy loss in our study is likely to be ecological and although we adjusted our analysis for as many factors as was possible, there might still have been some factors that follow a similar seasonality to temperature which were not included in our analysis.

In summary, we report a clear relationship between higher temperatures at conception and pregnancy loss in our sub-Saharan African setting. The effects of global warming will likely exacerbate the current reproductive health challenges faced by women in sub-Saharan Africa. Our study provides evidence for policymakers in South Africa and other countries in the sub-Saharan African region to give adequate attention to women’s reproductive health and pregnancy outcomes when drafting health policies in response to global warming.

## Supporting information

STROBE Checklist

## Data Availability

The data underlying this article are available (upon reasonable request) from the Africa Health Research Institute Data Repository.

https://data.ahri.org/index.php/home

## Acknowledgements

YM contributed to the study’s conception, design, statistical analysis, and drafting of the manuscript. FT contributed to the study’s conception and drafting of the manuscript. AT contributed to the study’s conception, design, acquisition of the temperature data from the South African Weather Services and drafting of the manuscript. All authors read and approved the final version of this manuscript. The authors are grateful to the study participants and the work and support of the fieldwork and database teams at AHRI. We would also like to thank the South African Weather Services for providing us access to their dataset. The corresponding author was supported with a postdoctoral fellowship under a National Institute of Health (NIH) grant (R01 HD084233). The Africa Health Research Institute’s Demographic Surveillance Information System and Population Intervention Programme is funded by the Wellcome Trust (201433/Z/16/Z), and the South African Population Research Infrastructure Network (funded by the South African Department of Science and Technology and hosted by the South African Medical Research Council). The funders played no role in designing the study; in the collection, analysis and interpretation of data; in the writing of the report; and in the decision to submit the article for publication. The content of this manuscript is solely the responsibility of the authors and does not necessarily represent the official views of the funding bodies.

## List of Abbreviations

AHRI: Africa Health Research Institute
95% CI: 95% Confidence interval
OR: Odds ratio
UNFCCC: United Nations Framework Convention on Climate Change

